# The Rapid Online Cognitive Assessment

**DOI:** 10.1101/2024.09.05.24313118

**Authors:** Calvin W. Howard, Amy Johnson, Joseph Peedicail, Marcus Ng

## Abstract

**INTRODUCTION:** Paper-based screening examinations are well-validated but minimally scalable. If a DCA replicate paper-based screening, it would improve scalability while benefiting from their extensive validation.

**METHODS:** We developed and evaluated the Rapid Online Cognitive Assessment (RoCA) against gold-standard paper-based tests in patients with a range of cognitive integrity (n = 46). Patient perception of the RoCA was also evaluated with post-examination survey.

**RESULTS:** The RoCA classifies patients similarly to gold standard paper-based tests, with a receiver operating characteristic area under the curve of 0.81 (95%CI 0.67-0.91, p < 0.001). It achieves a sensitivity of 0.94 (95%CI 0.80-1.0, p < 0.001). This was robust to multiple control analyses. 83% of patient respondents reported the RoCA as highly intuitive, with 95% perceiving it as adding value to their care.

**DISCUSSION:** The RoCA may act as a simple and highly scalable cognitive screen.

## Introduction

Current projections estimate 150 million dementia patients worldwide by 2050, with 57 million as of 2019.^1^ This causes considerable healthcare system strain, leaving a majority of patients undiagnosed.^2–5^ However, for the patients that do get a diagnosis, it often takes 3-years or longer from symptom onset.^2,3,6–8^ The next step, receiving an etiological diagnosis like Alzheimer Disease, requires even more time.^4,8^

Much research has previously focused on evaluating contributors to these problems.^9–13^ Frontline physicians report two key addressable issues: 1) the logistical difficulty of screening enough patients,^9,10^ and 2) variable comfort in diagnosing patients with dementia.^6,14^

Digital cognitive assessments (DCA) offer a promising solution to these issues.^15–17^ They provide high scalability, which addresses logistical difficulties, and can render expert-level diagnoses, which addresses the issue of diagnostic comfort. However, most DCAs have limitations preventing them from completely addressing these two issues.^18^

First and foremost is that these novel tests lack validation.^15^ DCAs often use completely new testing methods, diverging from the well-validated methods which made paper-based tests so valuable.^15–17,19,20^ Second, poor design choices often reduce accessibility.^16,17,21–25^ Common examples are burying test access deep within websites, requiring users to make accounts, or having patients use unfamiliar hardware.^16,17,21–25^ Lastly, most DCAs are not truly scalable.^16,17,21–25^ Despite being digital, they often require specific tablets, computers, on-site testing, or even expert test evaluators— these choices bottleneck patient access.^19,26,27^

The Rapid Online Cognitive Assessment (RoCA) is a DCA aiming to address these limitations. To stay consistent with well validated methods, it reproduces the screening results of gold-standard paper-based tests: Addenbrooke’s Cognitive Examination-3 (ACE-3) and the Montreal Cognitive Assessment (MoCA).^28–31^ Patient input directly guided its design to ensure accessibility, resulting in a short touchscreen-based drawing battery with automated convolutional neural-network based scoring.^32,33^ To provide scalability, it is entirely automated, remote, functions on all devices, and is available to roughly 75% of the globe.^34^

Finally, the RoCA aims specifically to act as a screening examination. For this reason, we prioritized its sensitivity.

We first make sure the RoCA’s underlying machine learning works well by assessing the accuracy of its neural network. Then, we compare the RoCA’s similarity to gold standard paper-based tests, evaluate its accessibility computationally and with patient input. Lastly, we provide the data-driven thresholds which maximize its sensitivity, optimizing its function as a screening examination.^35,36^

## Methods

### Ethics Statement

The study has been conducted in accordance with the ethical standards. This study was conducted in accordance with ethical standards as laid down in the 1964 Declaration of Helsinki and its later amendments. Approval was achieved by the Research Ethics Board of the Bannatyne Campus, University of Manitoba (#HS25666).

### Patients

Our study cohort enrolled patients from neurology clinics across the Health Sciences Centre, University of Manitoba (n = 46). Patients with and without cognitive complaints were recruited. Inclusion criterion was English fluency. Exclusion criteria were acute psychiatric disorder contributing to cognitive state, disability restricting ability to utilize screens, disability restricting ability to receive visual and auditory instructions, developmental delay, acute medical condition contributing to cognitive state, and specifically delirium. Patients indicating interest in clinical research were contacted by study team members via phone.

Interested patients were screened for inclusion and exclusion criteria and enrolled. At the first clinic visit, patients were again screened for inclusion/exclusion criteria by a physician. Patients or their caregivers provided written consent at the first clinic visit. Patients were recruited until sample size for statistical power was achieved.

### Study Design

Patients were tested in a quiet environment by a physician trained in cognitive examination. The RoCA was completed on a touchscreen tablet. The RoCA automatically administered instructions to the patient. It was completed without interference or prompting from the examiner. Caregivers were allowed to join but could not participate in the examination. RoCA responses were automatically scored and summated. The ACE-3 and MoCA examinations were administered and scored according to standard guidelines by a trained physician.^30,31^

### Cognitive Status Classification

A trained clinician administered a label of cognitive impairment based on established cutoffs for each test: 26/30 on the MoCA and 83/100 on the ACE-3.^30,31,37^

### Sample Size Calculation

Patients were enrolled based on sample size requirements defined by the Hansen and MacNeily formula.^38^ This formula is based on the area under the curve (AUC) of the receiver operating characteristic (ROC), and describes how sample size requirements vary with the AUC. Under relatively good performance, an AUC of 0.70, 80% statistical power is achieved with 16 positive and 16 negative cases. Under optimal performance with an AUC of 0.90, 80% statistical power is achieved with 2 positive and 2 negative cases. We conservatively aimed to recruit 16 positive and 16 negative cases.

### The RoCA

The RoCA is a self-administering cognitive screen which is compatible with smartphones, tablets, and personal computers. It relies upon devices having an internet connection to ensure all patients can access it regardless of hardware specifications or specific device.

The RoCA consists of 3 questions. Similar to previously described batteries, patients are asked to copy a line diagram of a cube, overlapping infinities, and perform a clock drawing.^32^ For each question, the patient has an unrestricted amount of time to answer. Each question’s instructions are provided via closed-captioned audio.

Instructions may be repeated up to 3 times, but no further assistance is provided. Questions are answered via touchscreen, although a keyboard and mouse may be used.

A correct cube drawing is worth 2 points, infinities 1 point, and a clock 5 points. Incorrect drawings are worth 0 points. The total possible score is 8 points.

### RoCA Deployment

The RoCA evaluation starts with the clinician (Figure 1). The clinician uses an administrative platform, from which they send tests and view results, to send an encrypted access link to a patient. Patients open this link to begin the RoCA. Links are specific to each given patient and inactivate after use. Upon completing the test, the results are encrypted and sent to a scoring server on a private subnet. The scoring server sends encrypted scores to an encrypted database on a private subnet. The administrative platform receives scores from this database, allowing the clinician to view results upon completion. The system is Health Information Privacy Protection Act compliant.

**Figure 1.**
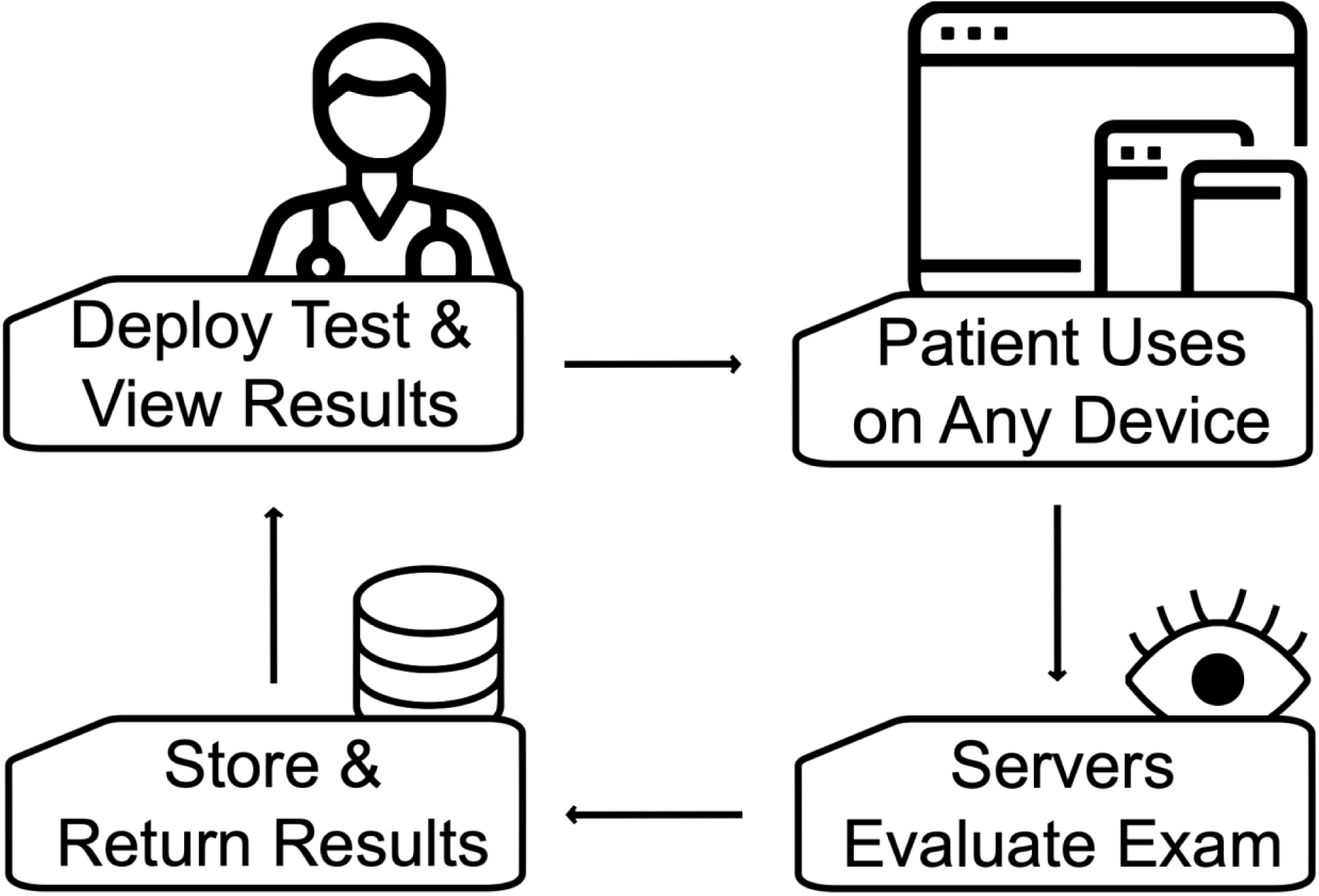
The RoCA deployment system. Clinicians begin the deployment system. They can use the administrative platform to deploy an access link to patients. Patients subsequently receive the access link and are then able to take the interactive test on any device via the internet. Once the patient completes the examination, their answers are passed to the servers which then evaluate the patient’s RoCA. The results are then stored in a database and are available for the patient’s clinician to see. Clinicians can view a patient’s results from the administrative platform.

### Patient Drawing Classification

Patient drawings are evaluated by the SketchNet, a convolutional neural network built specifically to evaluate RoCA inputs (Figure 2).^33,39^ Briefly, the SketchNet is a convolutional neural network using a SqueezeNet architecture.^40^ The particular architecture is composed of convolutional layers, fire modules, and finishes with a global average and SoftMax. This allows high degrees of accuracy while maintaining speed of classifications and a small overall size of the model. It was trained using transfer learning, and was pretrained on ImageNet, and subsequently trained on thousands of RoCA-specific drawings to evaluate cognitive test drawings with 97% accuracy.^39,40^

**Figure 2.**
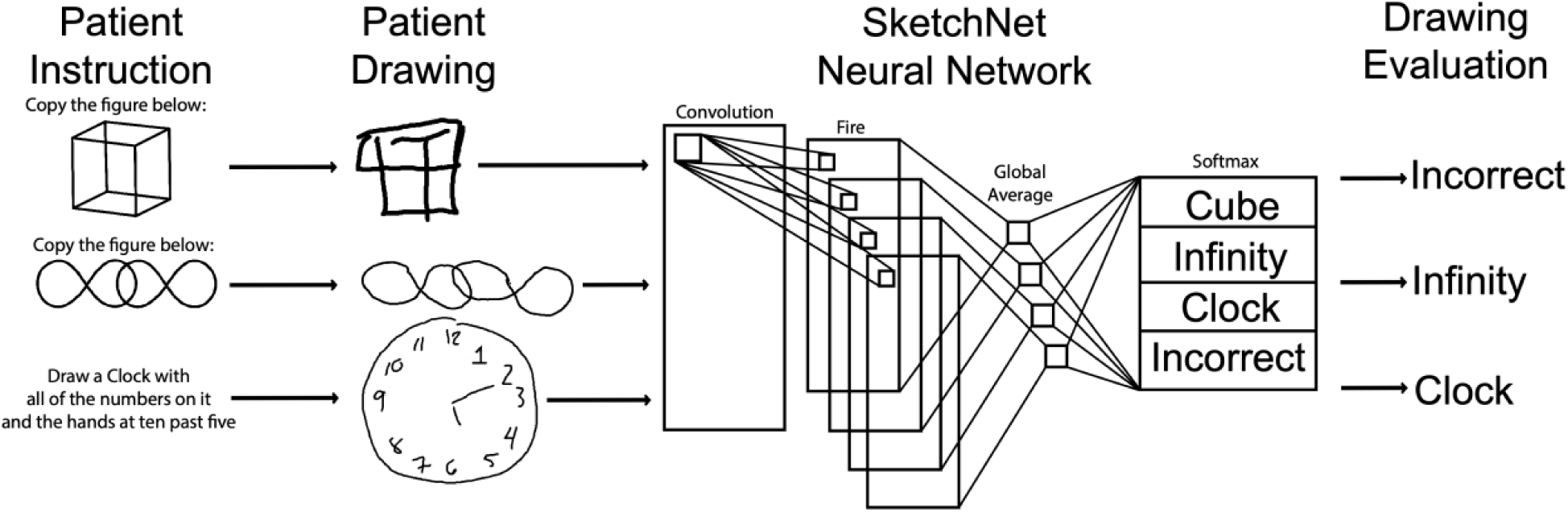
The RoCA drawing evaluation system. Patients receive audiovisual instructions asking them to draw three different images. The first two tasks are image copying tasks of a wire cube and overlapping infinities. The last question is to draw a clock face at ten-past-five. The patient drawings are then retrieved and preprocessed. After preprocessing, the images are then passed to the SketchNet, a convolutional neural network, to classify the images. The output of the SketchNet is the classification of the input image. The SketchNet is then applied to each image in the RoCA.

SketchNet-based classifications were compared to ground truth: assessment of drawings by a clinician trained in cognitive examinations. The drawings were scored according to established scoring guidelines.^30^

### Evaluation of Drawing Classification

The SketchNet is inherently a multiclass classifier, which yields complex classification evaluations. To simplify this, we condense classifications into correct versus incorrect. Confusion matrices were constructed from SketchNet classification outputs. Using the confusion matrix, we derived all classification metrics: accuracy, sensitivity, specificity, positive predictive value (PPV), and negative predictive value (NPV). This was repeated for each drawing.

### Comparison of Drawing Classification Against a Statistical Baseline

For a statistical baseline compare drawing classifications against, we derived a random classifier for each image. This random classifier represents performance at a random chance level. It is calculated by deriving a confusion matrix (true positives, true negatives, false positives, false negatives) under chance circumstances. The confusion matrix is thus the probability of selecting a given class multiplied by the probability of an image being a given class (Supplementary Equation 1). All classification metrics then follow from this confusion matrix. These provide the chance-level baseline.

To compare the classification of each drawing, we bootstrapped SketchNet classifications (n = 1000), and counted the number of times the bootstrap fell below chance level. This is the p-value. To compare the overall performance, we averaged performance across the RoCA and chance-level performance, then compared them with an independent T-test.

### Evaluation of Diagnostic Classification

To evaluate the reliability of RoCA patient classifications, we compared RoCA classification to ground-truth classification. Ground-truth classification was the classification adjudicated by the ACE-3 or MoCA in accordance with established guidelines.^30,31,37^ A receiver operating characteristic (ROC) was constructed, and its area under the curve (AUC) was calculated to measure diagnostic performance.

Youden’s Index was calculated to derive optimal classification threshold.^41^ Patient classifications were based upon this threshold, and these were then used to construct confusion matrices for the RoCA.

### Statistical Evaluation of RoCA Classification

To derive a statistical baseline for the RoCA, we again employ a random classifier equivalent to the RoCA. The confusion matrix is again generated using probabilities of selecting each class, as described above (Equation 1). The random-chance baseline for the AUC was chosen to be 0.50, in accordance with the literature.^38^

To compare the RoCA to these chance-level baselines, we again used the bootstrapping technique described above (n = 10 000).

### Classification Confidence

To evaluate the confidence of RoCA classifications, we estimate this directly with the confidence intervals derived from bootstrapping (n = 10 000).^42,43^ At all possible RoCA diagnostic thresholds, we derived the confusion matrices and classification metrics for each bootstrap. This allows observation of RoCA confidence across all possible thresholds. To make sure an appropriate threshold is chosen to optimize the RoCA for a screening examination, we focus on evaluating the sensitivity and NPV across all thresholds.

### Covariates Influencing RoCA Score

We aimed to identify clinical covariates which might be influencing RoCA scores. To do this, we collected several covariates: age, ethnicity, sex, educational status, employment status, and which paper-based exam they received. These covariates were then related to RoCA scores. This was done using a multivariate regression of all variables upon RoCA score.

We also assessed if any individual covariate compounded the effect of impaired cognition. This was done with a series of additional regressions. In these, the covariate, cognitive status, and their interaction were regressed upon RoCA score. This was done for each covariate.

### RoCA Usability and Patient Perception

A follow-up survey was sent to patients within six months of completing the RoCA. Patients responded to a battery of questions using both dichotomous (yes or no) questions and Likert scale questions. Likert scales were adjudicated such that 1 corresponded to very low, 2 was low, 3 was moderate, 4 was high, and 5 was very high.

### Statistics

All analyses were performed in Python. T-testing was performed with SciPy.^44^ Sci-Kit Learn was used for ROC construction.^45^ Regression analyses were performed with Statsmodels, and were performed with an ordinary least squares regression.^46^ Confidence intervals were calculated using bootstrapping with replacement (n = 1000).^42^

## Results

### Patient Characteristics

143 patients were assessed for eligibility. 79 patients declined enrollment. 7 did not meet inclusion criteria. 57 were enrolled, and 11 did not make their appointment. 46 completed the study. There were 16 patients with cognitive impairment and 30 patients without cognitive impairment. Patient demographics are available (Table 1).

**Table 1.** Patient demographics.

| Parameter | Overall |
| --- | --- |
| n | 46 |
| Age, mean (SD) | 49.1 (15.0) |
| Sex, n (%) |  |
| Female | 24 (52.2) |
| Male | 22 (47.8) |
| Educational Status, n (%) |  |
| Less than Secondary | 2 (4.3) |
| Post-Secondary | 8 (17.4) |
| Secondary | 36 (78.3) |
| Employment Status, n (%) |  |
| Employed | 29 (63.0) |
| Unemployed | 17 (37.0) |
| Ethnicity, n (%) |  |
| African | 1 (2.2) |
| Caucasian | 31 (67.4) |
| European | 1 (2.2) |
| Filipino | 3 (6.5) |
| Indian | 4 (8.7) |
| Indigenous | 6 (13.0) |
| Cognitive Status, n (%) |  |
| Impaired | 17 (37.0) |
| Intact | 29 (63.0) |
| Cognitive Exam, n (%) |  |
| ACE-3 | 35 (76.1) |
| MoCA | 11 (23.9) |

### The RoCA Can Accurately Evaluate Patient Drawings

We first evaluated how the RoCA evaluated patient drawings (Figure 3A-C). The RoCA classified 97% of cubes correctly (n = 44), 91% of infinities correctly (n = 42), and 98% of clocks correctly (n = 45).

**Figure 3.**
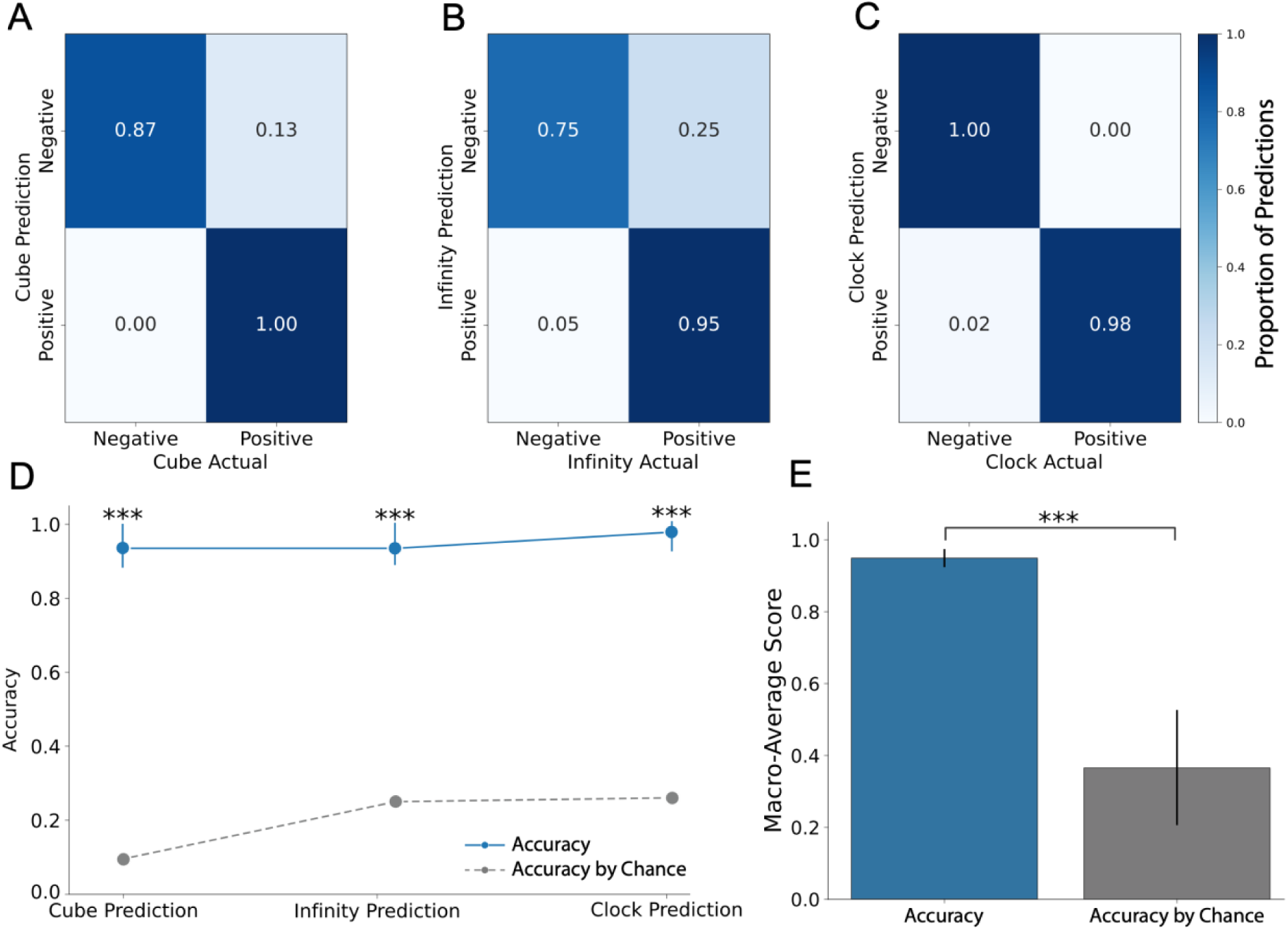
RoCA accurately classifies patient drawings. A) Confusion matrix for cube classification B) Confusion matrix for infinity classification. C) Confusion matrix for clock classification. D) Accuracy of the RoCA compared to the accuracy expected by chance, for all drawings. Bootstrapped confidence intervals were used to statistically compare the observed accuracy of the RoCA to the accuracy expected by chance. E) The overall accuracy of the RoCA is significantly higher than the accuracy expected by chance.

We next calculated the accuracy of the RoCA for each drawing individually (Figure 3D). We compared the accuracy of each drawing to its statistical baseline using bootstrapping, resampling the accuracy and counting the number of times it fell below the random classifier. Accuracy for the cube was 93% (95%CI 0.85-1.0, p < 0.001), the overlapping infinities was 94% (95%CI 0.87-1.0, p < 0.001), and the clock was 98% (95%CI 0.90-1.0, p< 0.001).

Finally, we derived the overall accuracy of the RoCA, across all drawings (Figure 3E). The RoCA had 95±3% accuracy across all drawings, which was higher than expected by chance (p < 0.001). Additional classification metrics are available (Supplementary Table 1)

### The RoCA Achieves Similar Diagnostic Fidelity to Gold-Standard Tests

Next, we compared the overall performance of the RoCA to the gold-standard tests (Figure 4). Each patient’s RoCA score was related to their diagnostic classification using an ROC compared to ACE-3 and MoCA classifications. The AUC was 0.81 (95%CI 0.67-0.91, p < 0.001). This was robust regardless of whether using the ACE-3 (AUC = 0.79, 95%CI 0.67-0.86, p < 0.001), or the MoCA (AUC = 1.0, 95%CI 1.0-1.0, p < 0.001). Subsequently, we found the optimal threshold for the RoCA was 7/8, according to Youden’s Index.^47^

**Figure 4.**
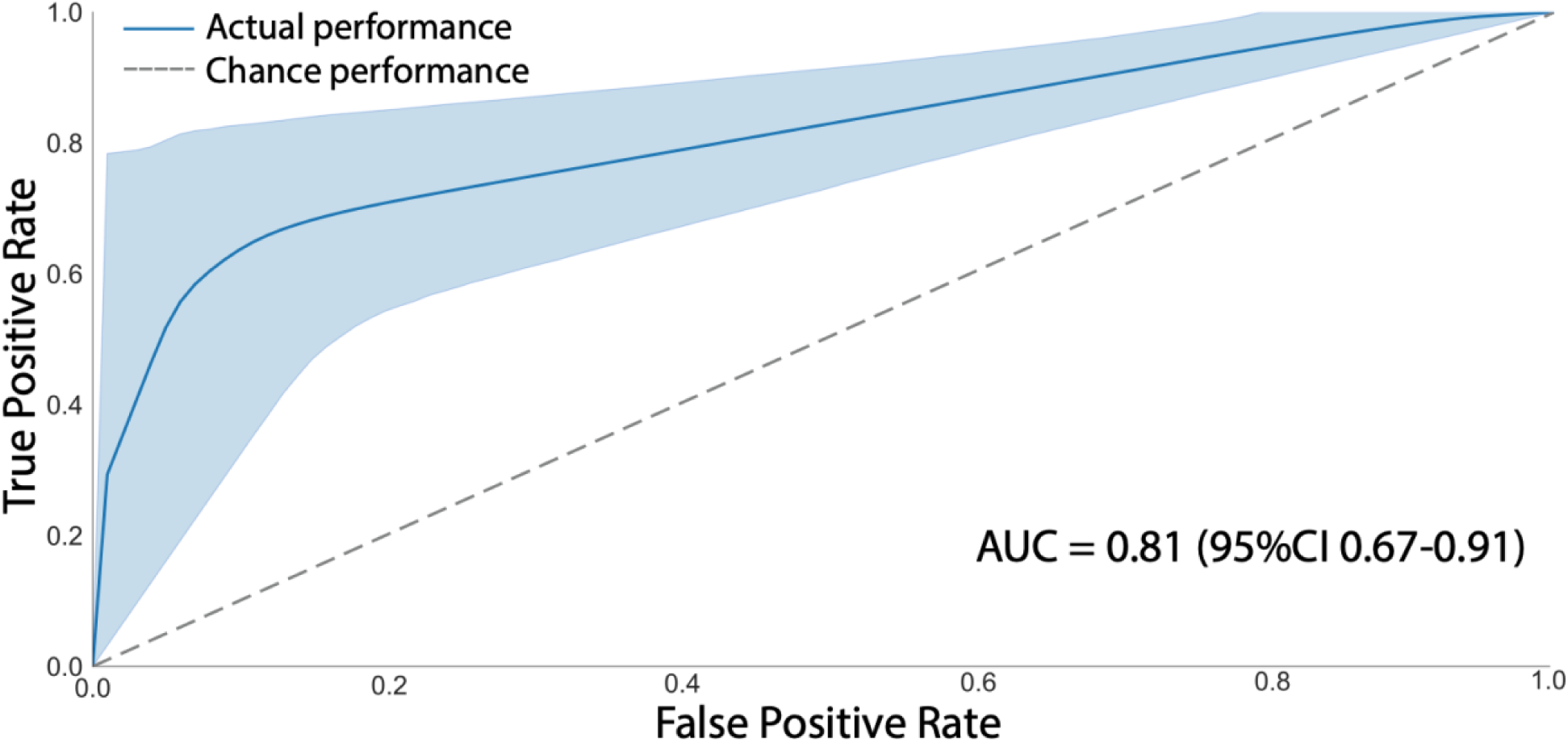
RoCA achieves high diagnostic performance compared to gold-standard paper-based test. ROC of the RoCA, compared to ACE-3 and MoCA (AUC = 0.81 (95%CI 0.67-0.91, p < 0.001). The ROC curve was calculated by bootstrapping the RoCA’s overall classification to derive confidence intervals and mean performance. The AUC presented is the mean AUC across all bootstraps. Shaded region represents 95% confidence interval. The point-estimate AUC, or the AUC without any bootstraps, is 0.85.

### The RoCA Achieves High Screening Performance

We next evaluated the RoCA’s ability to act as a screening examination. To do this, we evaluated the RoCA’s accuracy, sensitivity, and NPV. We began by developing a random classifier equivalent of the RoCA, which we used to derive the RoCA’s statistical baseline for comparison (Supplementary Figure 1). All screening metrics were expected to be low by chance, with an expected accuracy of 50%, sensitivity of 50%, and NPV of 63%.

Following this, we evaluated the actual RoCA’s screening performance. We began by calculating the confusion matrix of the actual RoCA, using the optimal threshold of 7/8 (Figure 5A). We then calculated the screening metrics for the RoCA (Figure 5B). At the optimal threshold, the RoCA has an accuracy of 0.76, which was better than expected by chance (95%CI 0.63-0.89, p < 0.001). It also achieved both superior and statistically significant sensitivity of 0.94 (95%CI 0.81-1.0, p < 0.001), and statistically significant NPV of 0.95 (95%CI 0.84-1.0, p < 0.001). Specificity and PPV were also calculated for completeness, although they are not directly related to screening ability (Supplementary Table 2)

**Figure 5.**
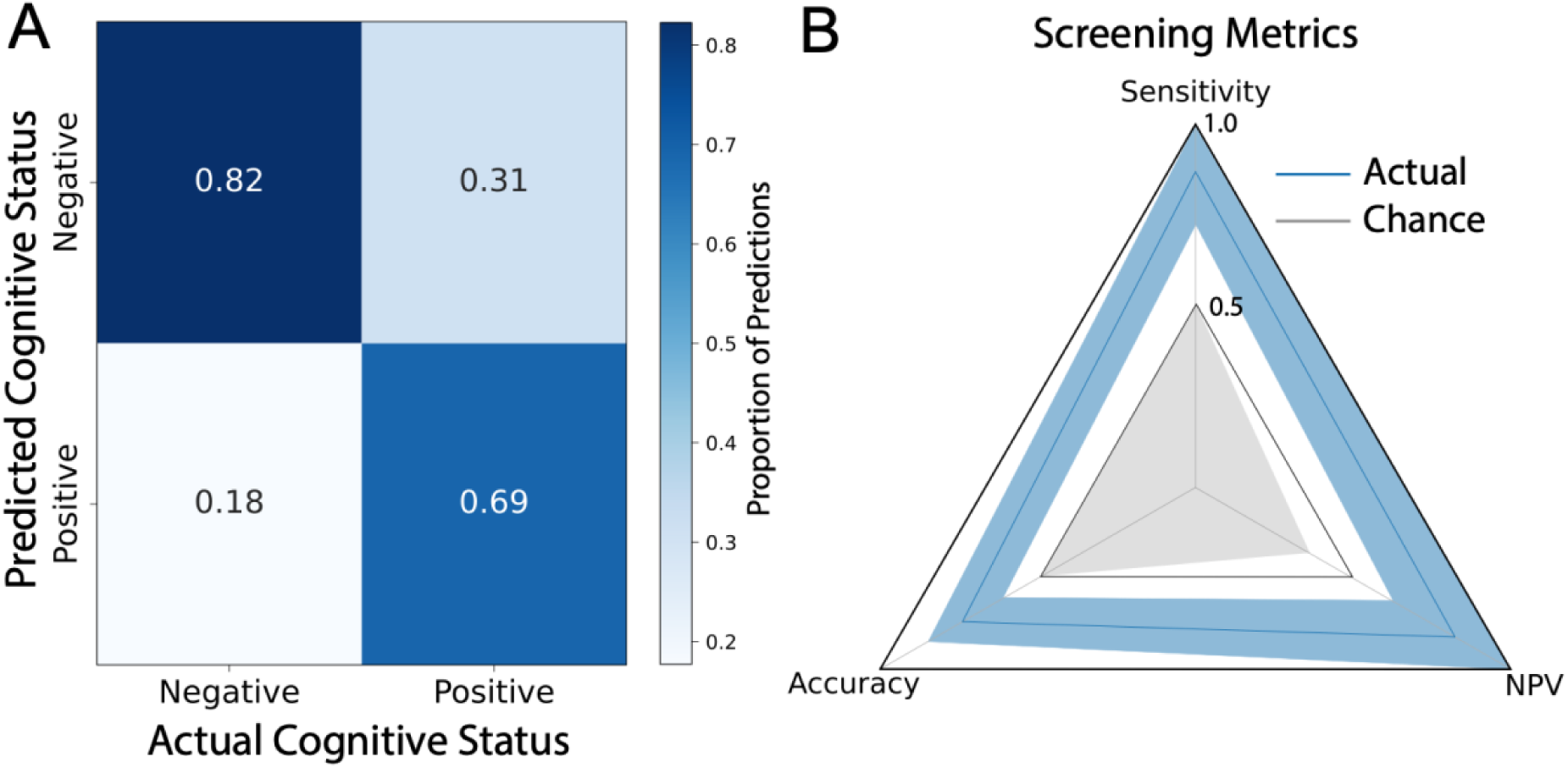
RoCA Achieves Excellent Screening Performance. A) Confusion matrix for classification of the RoCA at the threshold, normalized by predictions. True positive rate was 62% (n = 16), true negative rate was 95% (n = 19), false negative rate was 5% (n = 1), and false positive rate was 38% (n = 10). B) Screening metrics of the RoCA presented in a radar plot. Solid blue vertices represents the measured screening metric, with shaded blue edges marking the 95% confidence intervals. The grey interior represents the expected performance by random chance. The RoCA achieved an excellent sensitivity of 0.94 (95%CI 0.63-0.89, p < 0.001) and an excellent NPV of 0.95 (95%CI 0.84-1.0, p < 0.001). Accuracy is also presented, although it is not purely a screening metric, and was better than expected by chance at 0.76 (95%CI 0.63-0.89, p < 0.001).

### RoCA Diagnostic Confidence

Next, we ensured the chosen RoCA threshold is the optimal threshold for screening. Youden’s J, calculated using the ROC, balances sensitivity and specificity, and therefore will not necessarily result in the optimal screening threshold. To search for the optimal screening threshold, we calculated the confidence of sensitivity and NPV across all potential scores (Figure 6). We found the threshold of 7/8, identified by the AUC, also was the optimal screening threshold. It simultaneously maximized the sensitivity (0.94) and NPV (0.95), while also optimizing the confidence interval for sensitivity (95%CI 0.81-1.0) and NPV (95%CI 0.84-1.0). Specificity and PPV were also calculated for completeness and are available, although these are not optimized in screening examinations (Supplementary Figure 2).

**Figure 6.**
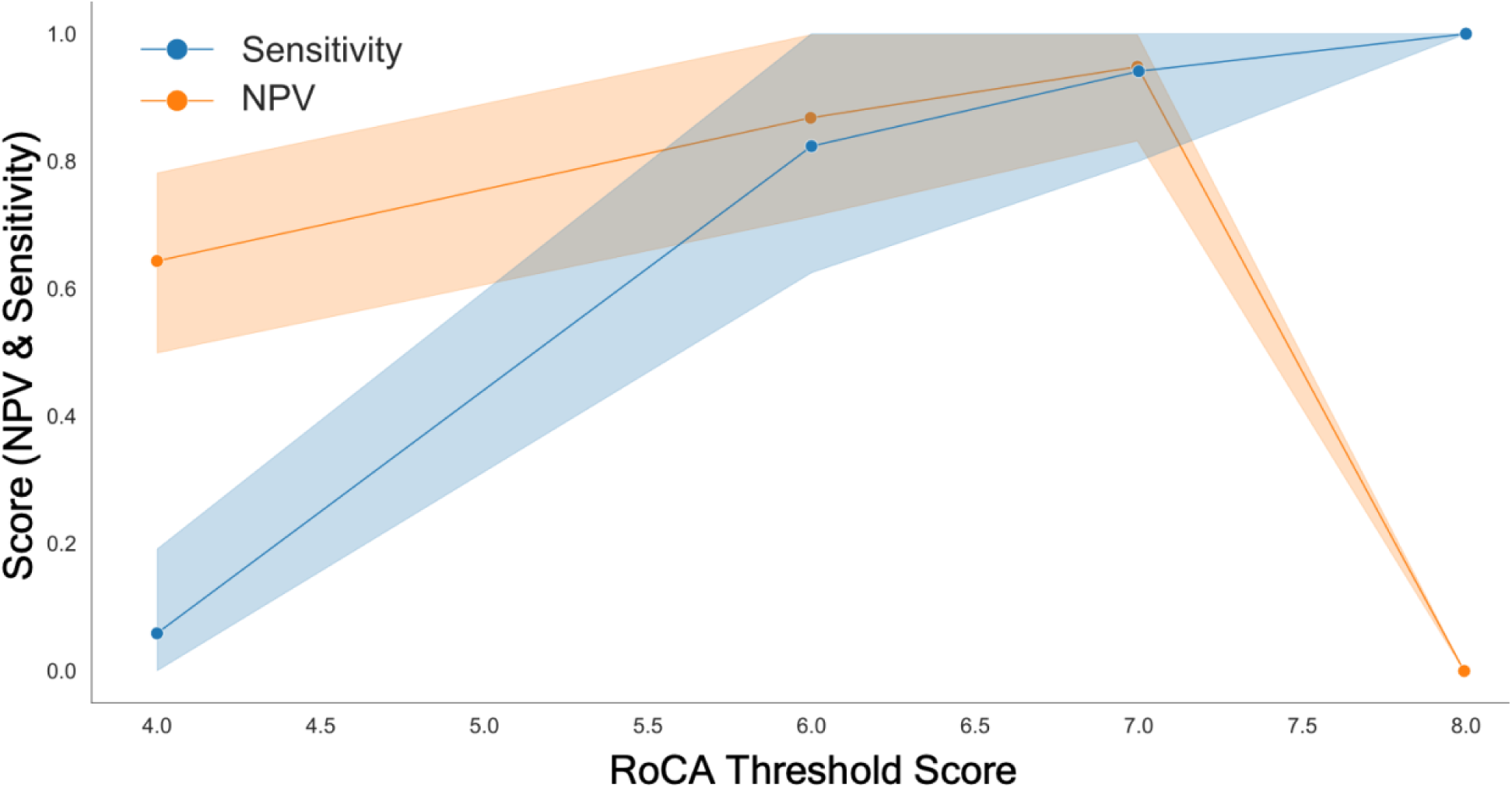
At the optimal threshold of 7/8, the RoCA optimizes both sensitivity and NPV. At 7/8, the values of sensitivity and NPV are maximized with minimization of their uncertainty. At this point, sensitivity is also highly confident (sensitivity = 0.94, 95%CI 0.80-1.0). NPV is also maximized (NPV = 0.80, 95%CI 0.83-0.95). Shaded regions represent 95% confidence intervals derived from bootstrapping (n = 10 000). Points represent the estimated sensitivity and NPV value without bootstrapping.

### Accessibility - The RoCA Score is Not Influenced by Patient Demographics

We next evaluated if any patient factors may be influencing RoCA performance. We first performed one multivariate regression to evaluate the relationship of all demographic variables upon RoCA score (Supplementary Figure 2). However, only cognitive status had a significant association with RoCA score (β = 1.07, p < 0.001). No other patient factor was related.

We also performed a series of multivariate regressions for each covariate, assessing if patient factors might compound the effects of impaired cognition (Supplementary Figure 3). Again, we found only cognitive status was significantly associated with RoCA scores.

Lastly, we evaluated the time investment required to complete the RoCA. We found the RoCA takes roughly two and a half minutes (148 ± 34 seconds).

### Accessibility – Patients Find the RoCA Accessible

Finally, we evaluated accessibility as reported by patients. We did this using post-RoCA survey responses wherein patients answered questions specifically regarding the accessibility of the RoCA (Figure 7). 19 of 46 patients responded (60.2 ± 18 years old). Cognitive status of respondents was unknown given anonymous nature of the survey.

**Figure 7.**
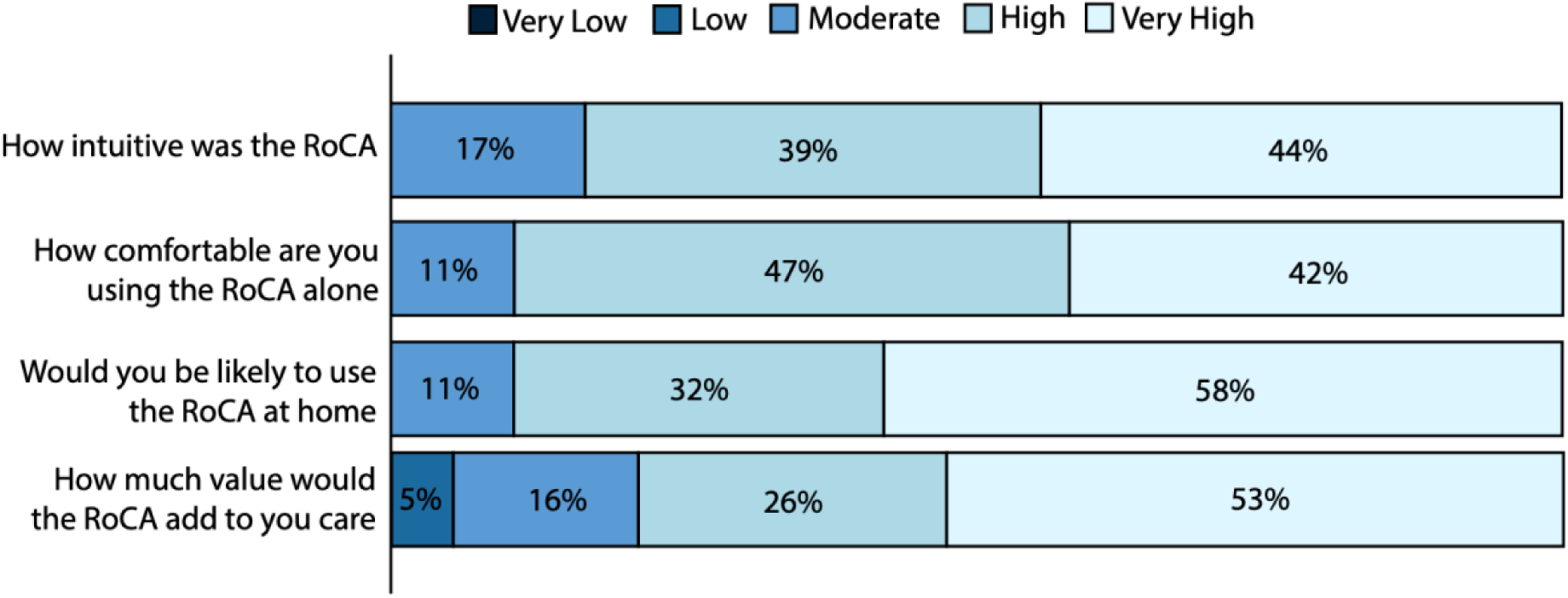
Patients find the RoCA accessible. Survey responses from 19 patients post-RoCA (60.2 ± 18 years old). Barplot of patient responses to Likert Scale questions. 83% of respondents reported the RoCA as at least highly intuitive. 89% of respondents reported they would be at least highly comfortable using the RoCA alone. 90% of respondents reported they would be at least highly likely to use the test at home. 79% of respondents reported the RoCA would add at least a high degree of value to their care.

Likert scales were used to derive an evaluation of overall patient evaluation of the RoCA. 83% of respondents reported the RoCA as highly or very highly intuitive. 89% of respondents reported high or very high comfort in using the RoCA. 90% of respondents reported they would be highly or very highly confident using the test alone. 79% of respondents reported the RoCA would add a high or very high degree of value to their medical care.

We also asked patients a series of yes or no questions regarding other aspects of the RoCA (Supplementary Table 3). Among them, we found 100% of patients would want to take the test prior to appointments to discuss results, and 100% of patients would trust the RoCA’s results. Most surprisingly, we found 55% of patients made appreciable lifestyle changes after the RoCA. These changes specifically included starting cognitive exercises, beginning a dementia-friendly diet, starting physical exercise, or doing financial planning.

## Discussion

### Interpretation of Results

We find the RoCA performs highly. It can accurately evaluate complex cognitive examination drawings and subsequently use them to sensitively screen for cognitive impairment. Importantly, it does this in-line with established paper-based tests. Further, we find the RoCA is a highly accessible screen, being unaffected by patient factors and well regarded by patients. This combined with the RoCA’s cloud-based platform allows it to act as a sensitive, accessible, and scalable digital cognitive screen.

### Role as a Screening Exam

The first limitation of DCAs is their lack of validity compared to paper-based screens.^15–17,19,20^ The RoCA specifically aimed to classify patients similarly to these paper-based screens, aiming to act as a digital surrogate for them.^28–31,48–50^ We find the RoCA rules out (screens) cognitively healthy patients which would have been similarly ruled out by standard paper-based tests.

The primary benefit of the RoCA is triaging patients for further examination. It can prioritize at-risk patients while offloading the cognitively healthy for routine observation. This process eliminates the proportion of true negatives from the population which go on for subsequent assessment, thereby increasing the positive and negative predictive values of any further evaluation.^51^ For these reasons, the RoCA is best used in a two-part system wherein the first test prioritizes sensitivity while the second prioritizes specificity. For example, the RoCA could be followed by our other full length diagnostic test, the Autonomous Cognitive Examination.^52,53^

### Limitations

This study and the RoCA are not without limitations. First, this study does not focus on individual etiologies causing dementia, but focuses on identifying cognitive impairment as a whole. Thus, it is possible that etiologies presenting in different cognitive domains may result in variable RoCA performance. However, before specializing into the evaluation of different etiologies, it is critical to accurately screen cognitive impairment itself. This will help the RoCA generalize across disorders causing cognitive impairment.

There are limitations to the RoCA. First, the RoCA is hardware dependent. For patients without touchscreens, they may have difficulty in generating high quality drawings, which may hinder performance. However, the RoCA was specifically trained on a dataset mixing drawing generated from touchscreens, mice, and stylets to specifically offset this risk. Beyond this, the RoCA is internet connection dependent. However, we have ensured this is compatible with smartphones to leverage their inherent internet connection.

## Data Availability

Data are available upon reasonable request.

## Acknowledgements/Conflicts/Funding Sources/Consent Statement

The authors would like to acknowledge Rachel Elizabeth Bethune Howard for support of the research.

Author CH is part of CogNet Inc., a company provided cognitive testing to rural Canadians.

Author AJ has no disclosures.

Author JP has no disclosures.

Author MN has no disclosures.

There is no funding to disclose.

Subjects were recruited from the neurology and neuropsychology clinics, and all signed Institutional Review Board-approved consent forms. Substitute decision makers were included in the consent process of cognitively impaired patients. This work was conducted between 2022 and 2024.

