## supplements for "The Rapid Online Cognitive Assessment"

$$P(prediction) = P(chose\ correct\ class) * P(is\ correct\ class)$$

Supplementary Equation 1. **Calculation of Random Classifier Confusion Matrix Components.** The probability that the random classifier chooses the correct class is the prevalence of that class amongst all classification choices. The probability that this same class is chosen is the prevalence of the observed class within the dataset. This allows each part of the confusion matrix to be calculated.

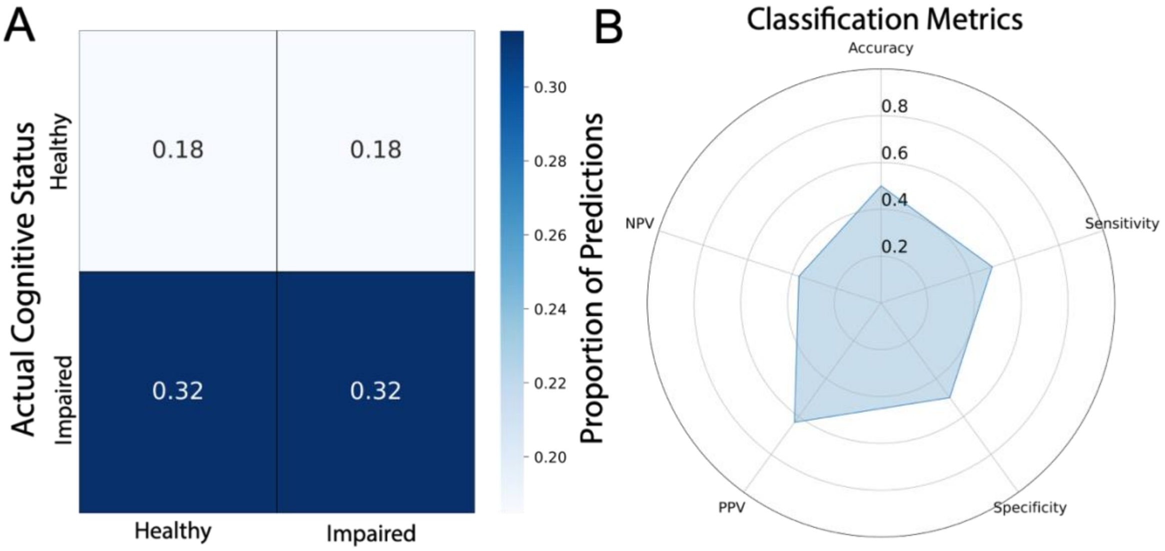


Supplementary Figure 1. **Random Classifier for RoCA Drawings.** A) Confusion matrix for random classifier. 18% of classifications will identify true negatives, 32% will identify true positives, 18% will identify false negatives, and 32% will be false positives. B) Radar plot of random classifier classification metrics (Accuracy = 0.50, Sensitivity = 0.50, Specificity = 0.50, PPV = 0.63, NPV = 0.37).

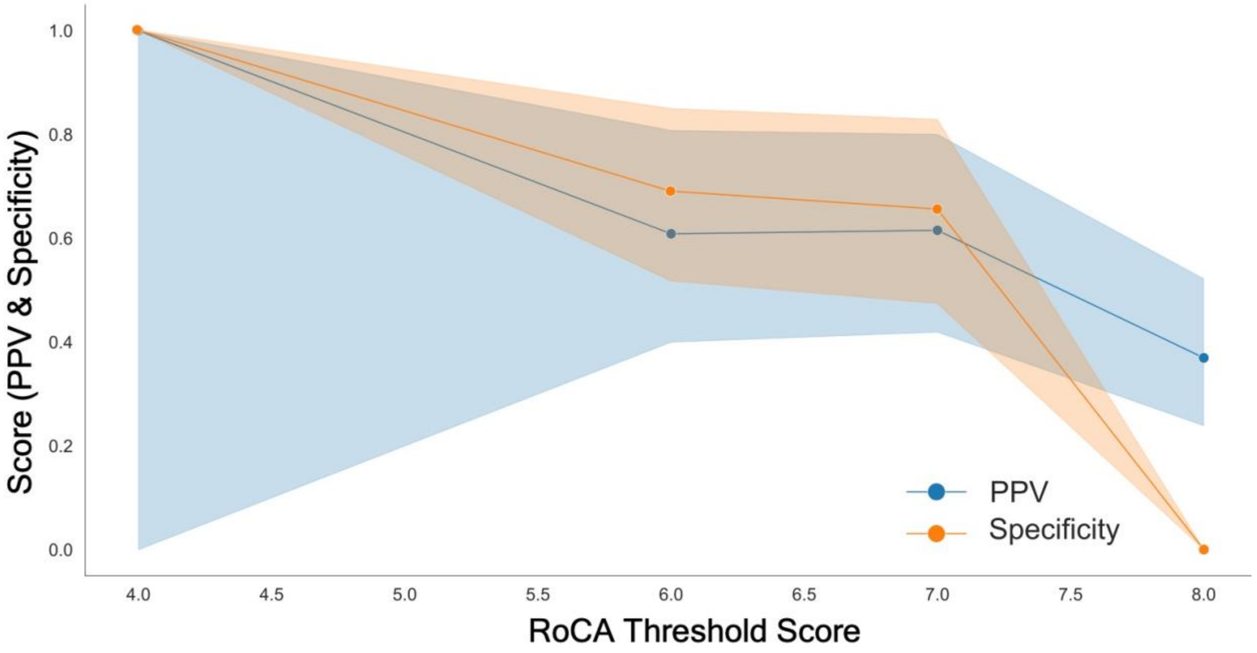


Supplementary Figure 2. **Certainty of sensitivity and NPV across RoCA scores.** Specificity confidence is maximized at a threshold of 4/8 points (specificity = 1.0, 95%CI 1.0-1.0). However, while point estimates of PPV at this threshold are 1.0, the certainty of the PPV is (95%CI 0.0-1.0). Shaded regions represent 95% confidence intervals derived from bootstrapping (n = 10 000). Points represent the estimated sensitivity and NPV value without bootstrapping.

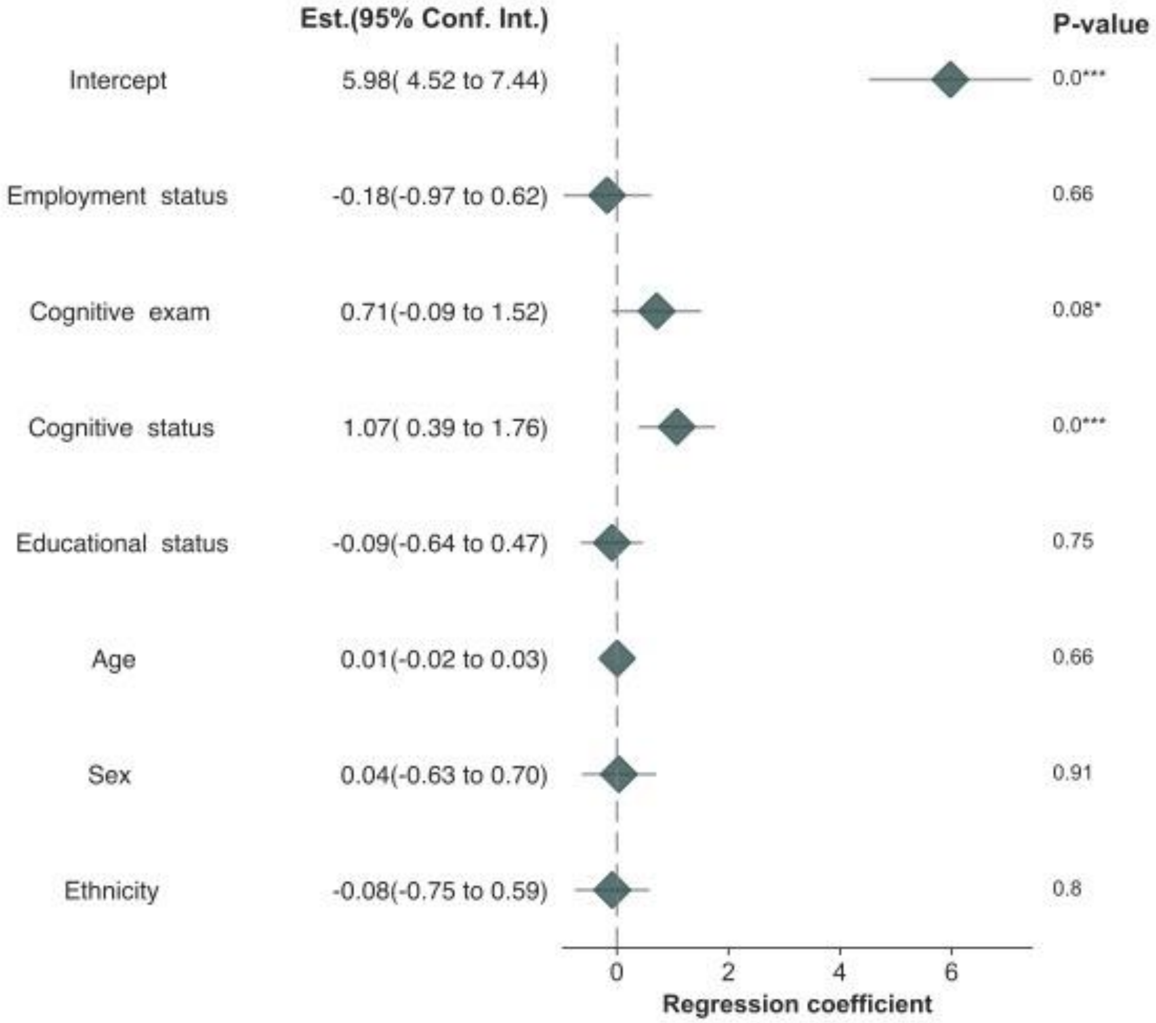


Supplementary Figure 3. **No covariate except for cognitive status is related to RoCA scores**. The entire multivariate regression is presented as a forest plot, with coefficients visualized as diamonds and their 95% confidence intervals as bars. P-values for each coefficient are shown at the right. Coefficients are on the scale of the RoCA, relating their impact directly to RoCA score.

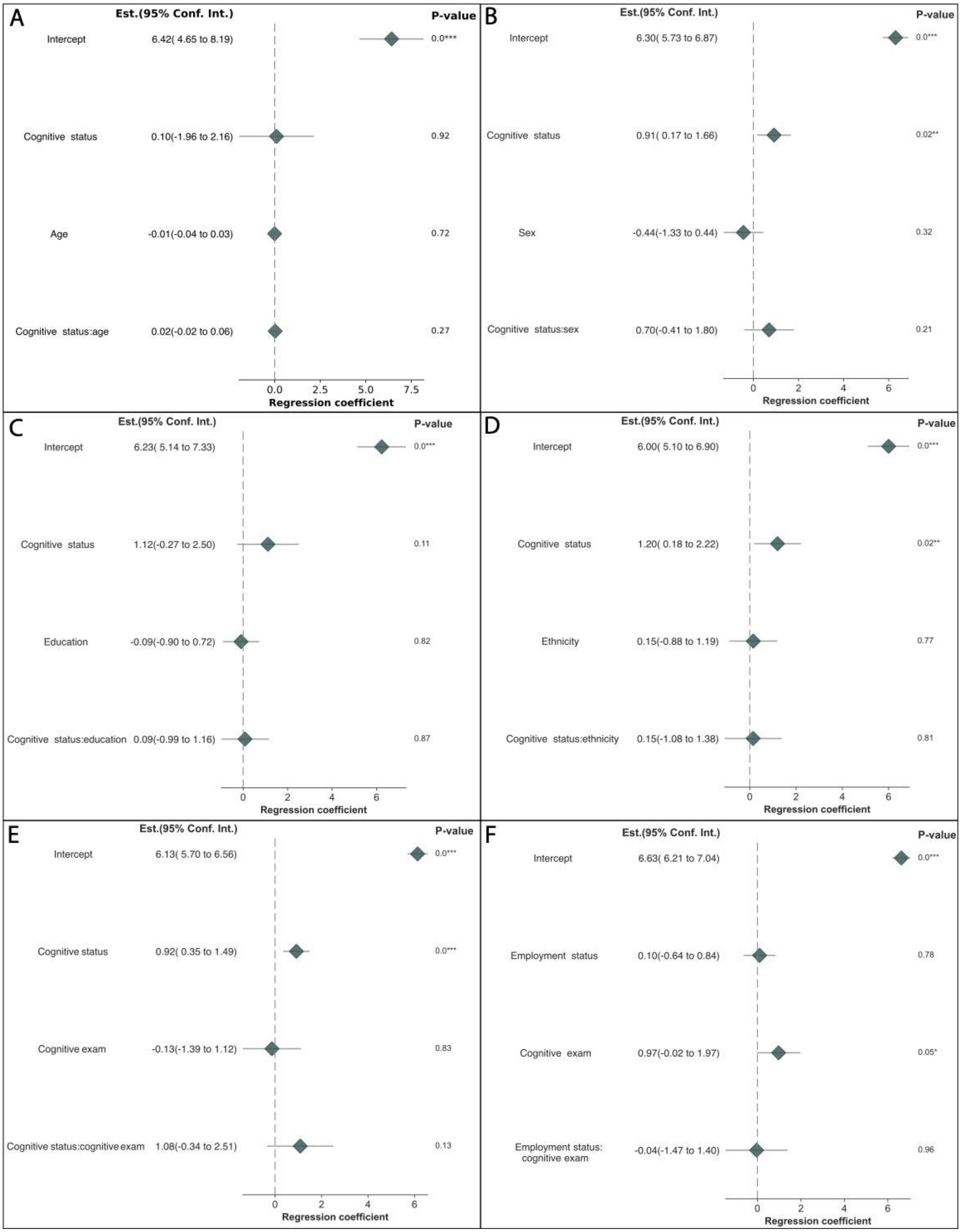


Supplementary Figure 4. **No covariate is related to RoCA scores aside from cognitive status.** A) Age after controlling for cognitive status and potential interactions. B) Sex after controlling for cognitive status and potential interactions. C) Education after controlling for cognitive status and potential interactions. D) Ethnicity after controlling for cognitive status and potential interactions. E) Cognitive examination type after controlling for cognitive status and potential interactions. F) Employment status after controlling for cognitive status and potential interactions. Each multivariate regression is presented as a forest plot, with coefficients reported as estimates with 95% confidence intervals. The coefficients and confidence intervals are forest plotted. P-values for each coefficient are shown at the right. Unstandardized coefficients are presented to ensure directly interpretability and application in relation to RoCA scores.

Supplementary Table 1. **Drawing Classification Metrics.**

| Drawing | Accuracy | Sensitivity | Specificity | Positive Predictive Value | Negative Predictive Value |
|---|---|---|---|---|---|
| Cube | 0.93 | 1.0 | 0.87 | 0.88 | 1.0 |
| Infinity | 0.94 | 0.95 | 0.75 | 0.98 | 0.60 |
| Clock | 0.98 | 0.98 | 0.75 | 0.98 | 0.02 |
| Overall | 0.95 | 0.95 | 0.70 | 0.98 | 0.87 |

Supplementary Table 2. **RoCA Classification Metrics.**

| | Accuracy | Sensitivity | Specificity | Positive Predictive Value | Negative Predictive Value |
|---|---|---|---|---|---|
| RoCA | 0.76 | 0.94 | 0.66 | 0.62 | 0.95 |

Supplementary Table 3. **Survey Responses.**

| Question | Yes | No |
|---|---|---|
| Would Like to Take Test Prior to Appointments | 100% | 0% |
| Would Recommend to Others | 100% | 0% |
| Would Add to Care | 100% | 0% |
| Taking Remotely Would Alleviate Stress of In-Clinic Exams | 95% | 5% |
| Made Lifestyle Changes as Result of Test | 55% | 45% |
| Would Trust the Results of the Test | 100% | 0% |
